# Examining the Evolution and Drivers of COVID-19 Transmission Waves in Ghana, 2020 – 2022

**DOI:** 10.1101/2023.07.17.23292790

**Authors:** Christopher Sunkwa Tamal, Sally-Ann Ohene, Rafiq Nii Okine, Michael Adjabeng, Argata Guracha Guyo, Fred Osei-Sarpong, Patrick Avevor, Ama Akyampoma Owusu-Asare, Franklin Asiedu-Bekoe, Gideon Kwarteng, Patrick Kuma-Aboagye, Francis Chisaka Kasolo

**Affiliations:** World Health Organization, Accra, Ghana; Ghana Health Service, Accra, Ghana

**Author notes:** Corresponding author: Christopher Sunkwa Tamal.

**Keywords:** COVID-19, transmission waves, lockdown, effective reproduction number, Ghana

## Abstract

**Background:** Ghana reported the first COVID-19 cases on 12 March 2020. Response actions were rolled out along seven thematic pillars to limit the importation, detect and contain the virus, effectively manage cases, ensure effective coordination and maintain essential services. A whole-of-government and whole-of-society approach was adopted for the response. The government instituted restriction measures at various stages of the response to contain the pandemic or limit the impact of the pandemic on the health, social and economic wellbeing of the citizens. Four distinct transmission waves were recorded within the first 2 years of the pandemic. The study examined the key drivers of the major waves.

**Methods:** A descriptive analysis of the pandemic from March 2020 to March 2022 was conducted using data reported through the country’s COVID-19 surveillance platforms. All RT-PCR confirmed cases reported from the 16 administrative regions over the two-year period were analysed. The effective reproduction number was computed using a model developed by Cori and colleagues.

**Results:** A total of 160,761 cases with 99.1% (159,227) recoveries or discharges were reported as of 12 March 2022. The Greater Accra Region reported 56.3% of the confirmed cases. Within the period, 1,445 deaths (CFR= 0.9%) were reported. Approximately 2.3 million tests (76,774 per million population) tests were conducted with a cumulative test positivity rate of 6.8%. COVID-19 vaccination was enrolled a year after the first cases were reported and 21.3% of the target population was fully vaccinated as of 12 March 2022. Ghana recorded four major COVID-19 transmission waves characterized mainly by variants of concern and sub-optimal adherence to the public health and social measures.

**Conclusion:** Scaling up and enhancing community acceptance of COVID-19 vaccination as well optimizing the current surveillance and response systems are essential is sustaining the current gains and limiting the emergence of new variants of concern.

## Background

The first two cases of COVID-19 in Ghana were recorded on 12 March 2020 (1,2), a day after WHO declared the disease as a pandemic. Countries were urged to adopt a whole-of-government and whole-of-society approach in the response to the disease while being mindful of economic and social disruptions and as well as respecting human rights in the quest to protect lives (3). At the outset, researchers postulated that the impact of COVID-19 on the Africa continent may be devastating owing to its fragile health systems (4).

Prior to the confirmation of the first COVID-19 cases in the country, the country had undertaken several preparedness activities including training of healthcare workers, media engagement, public sensitization, and stakeholder engagement (2). The country activated its response plans to contain the disease spread.

Ghana’s National Strategic Response Plan (NSRP) was built around six objectives: (i) Limit and stop the importation, detect and contain the virus; (ii) slow down and manage community spread; (iii) provide adequate medical and psychosocial care for COVID-19 cases; (iv) strengthen governance, coordination and accountability of COVID-19 response; (v) minimise impact of COVID-19 on social and economic life; and (vi) increase domestic capacity and self-reliance including building and strengthening capacity for health research and Innovations (5).

Response actions were organized and delivered around seven pillars namely Governance and Coordination, Containment and Case Management, Risk Communication and Social Mobilization, Testing and Laboratory Services, Points of Entry, Research and Innovation, and maintaining essential health services and systems (5).

Within two years of response, the country recorded four distinct major transmission waves. These waves had different impact on the health system as well as the wellbeing of the population. There is the need to examine the evolution and key drivers of the epidemiological trajectory and waves of the pandemic in the country in relation to the response actions taken. This will provide information for stakeholders and to intensify or redefine the response actions. It will also provide a framework for pandemic preparedness and response in the country.

The main objective of this study was to examine the key drivers which characterized the major transmission waves with the specific objectives to:

- Describe the trend of confirmed COVID-19 cases within the two years of the pandemic
- Examine the levels of the community transmission of COVID-19 in the country.
- Describe the measures taken by government to address the pandemic

## Methods

### 3.1 Study design

This is a descriptive study of COVID-19 pandemic in Ghana. The study described the trajectory of the pandemic in the first two years of the pandemic in juxtaposition with public health and social measures adopted in response to the pandemic and the variants of concern (VOCs).

### 3.2 Study setting

Ghana is in the West African sub-region. It has 16 administrative regions and 260 districts. Ten regional hospitals serve as referral centres for tertiary care. Each regional hospital has a designated area for the holding and managing mild to moderate COVID-19 cases. There are five teaching hospitals which provide advanced health care services including management of severe and critical cases of COVID-19. The teaching hospitals are strategically located to serve the northern, middle, and southern zones of the country. A specialized hospital, the Ghana Infectious Disease Centre (GIDC), was built during the COVID-19 pandemic to augment the other health facilities in the country.

Initially, two major laboratories, the Noguchi Memorial Institute for Medical Research (NMIMR) and the Kumasi Centre for Collaborative Research in Tropical Medicine (KCCR), were the only referral laboratories for PCR testing in the country (6). By the end of December 2020, over 20 public and private testing facilities were conducting RT-PCR COVID-19 testing.

### 3.3 Data collection and reporting

All data were extracted from the Ghana Health Service website for COVID-19 (7). The data were validated from the Disease Surveillance Department of the Ghana Health Service. Only aggregated data are available on the website. At the service delivery points, data on cases (suspected, probable, and confirmed) and contacts were captured electronically onto the Surveillance, Outbreak Response Management and Analysis System (SORMAS). The case-based data is transmitted in real-time through the levels of the health system (from health facility to national level) where data can be visualized and/or accessed depending on the level of access granted to users. Samples of cases and contacts were also received at the testing laboratories using a barcode system and the results transmitted to the specific facility or districts which submitted the samples. All confirmed cases were summarized and reported on the website.

### 3.4 Case Detection and Laboratory Diagnosis

Cases are confirmed using Real-time reverse transcriptase Polymerase Chain Reaction (RT-PCR). During the first wave of COVID-19 in Ghana, enhanced surveillance was employed to capture all suspected cases. By this approach, a 2-kilometer radius was mapped for every confirmed case and samples of persons living within the enclave collected for testing to establish the extent of community transmission (8).

The NMIMR subsequently developed and validated a pooling testing strategy, a well-known cost-effective technique particularly useful in public health surveillance of infectious diseases (9).

To augment the testing capacity particularly at the sub-national level, the GeneXpert platform was introduced as a point-of-care test mainly for symptomatic patients (6). This platform was operationalized in all the 16 regions of the country. On the Ghana Service website, only cases confirmed by PCR were reported.

### 3.5 Data Analysis

All the data were extracted from the Ghana Health Service Website for COVID-19. New cases for a particular day were computed by subtracting the cumulative confirmed cases on that day from the cumulative cases of the previous day. Test positivity rate was calculated by dividing the confirmed cases by the number of tests conducted for the same period expressed as a percentage. The number of deaths reported was calculated by subtracting the sum of recoveries and active cases from the total confirmed cases reported over the same period.

The time-dependent effective reproductive number (R_t_) was estimated using a WHO-recommended model to describe the level of ‘infectiousness and transmissibility’ of COVID-19 in the country (10).

R_t_ is the average number of secondary cases that each infected individual would infect if the conditions remained as they were at time t. The R_t_ was estimated using the formula 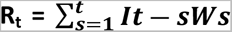 where

t = time step; *I*_t_ = Total infectiousness at a given time (t); s = time since the infection of the case; w=weighted infectivity/infectivity profile.

This formula was modelled into a Microsoft Excel sheet developed by the author (10). The parameters required for the model included the mean and standard deviation of the Serial Interval (SI) (time between symptom onset in primary case and symptom onset in secondary case), time step choice and prior distribution (mean and standard deviation). From a systematic and meta-analysis (11), the mean (Standard Deviation) SI was set at 5.2 (±0.52) and the prior probability set at mean = 5 and Standard Deviation = 5. The length of time step was set at 7 days for estimates at the end of 7-day period and 1 for the estimates to be performed every day. A good indicator of a slowing or declining epidemic is to maintain (R_t_) < 1 for at least 14 days (in the context of COVID-19).

Test positivity rate was used to define the level of community transmission. The categorization of the level of community transmission was done according to guidelines (12). Test positivity rate <5% for a two-week period was described as low community transmission, 5 – 20% was described as high community transmission while >20% was described as very high community transmission.

### 3.6 Ethical considerations

All the data set used in study were aggregate data. There were no patient identifiers which could be linked to any individual person or family. All data reported are part of the routine reporting system. Permission to use the data was obtained from the Ghana Health Service.

## Results

### Trend of confirmed COVID-19 cases within the two years of the pandemic

The 12 of March 2022 marks exactly two years since the first two COVID-19 cases were confirmed in Ghana. As of 12 March 2022, a total of 160,761 confirmed cases of COVID-19 had been reported in the country (Table 1). The cumulative number of recoveries and discharges was 159,227 (recovery rate= 99.05%). The country recorded four distinct major waves since the outbreak. Two minor waves were also recorded. The first was a minor wave which was recorded from the second week of April to the first week of May 2020 (Figure 1). This was followed by a major wave which lasted from the first week of June 2020 to the end of July 2020. A total of 29,192 cases were reported in the first major wave. Again, there was a minor wave from the third week of October to the second week of November 2020. The second major wave was recorded from the first week of January 2021 to the second week of March 2021 in which 34,432 COVID-19 cases were reported. The third major wave lasted from the fourth week of June 2021 to second week of October 2021 with 33,725 cases reported. The fourth major wave spanned from the second week of December 2021 to the end of February 2022 (Figure 1) and a total of 29,192 cases were reported.

**Figure 1:**
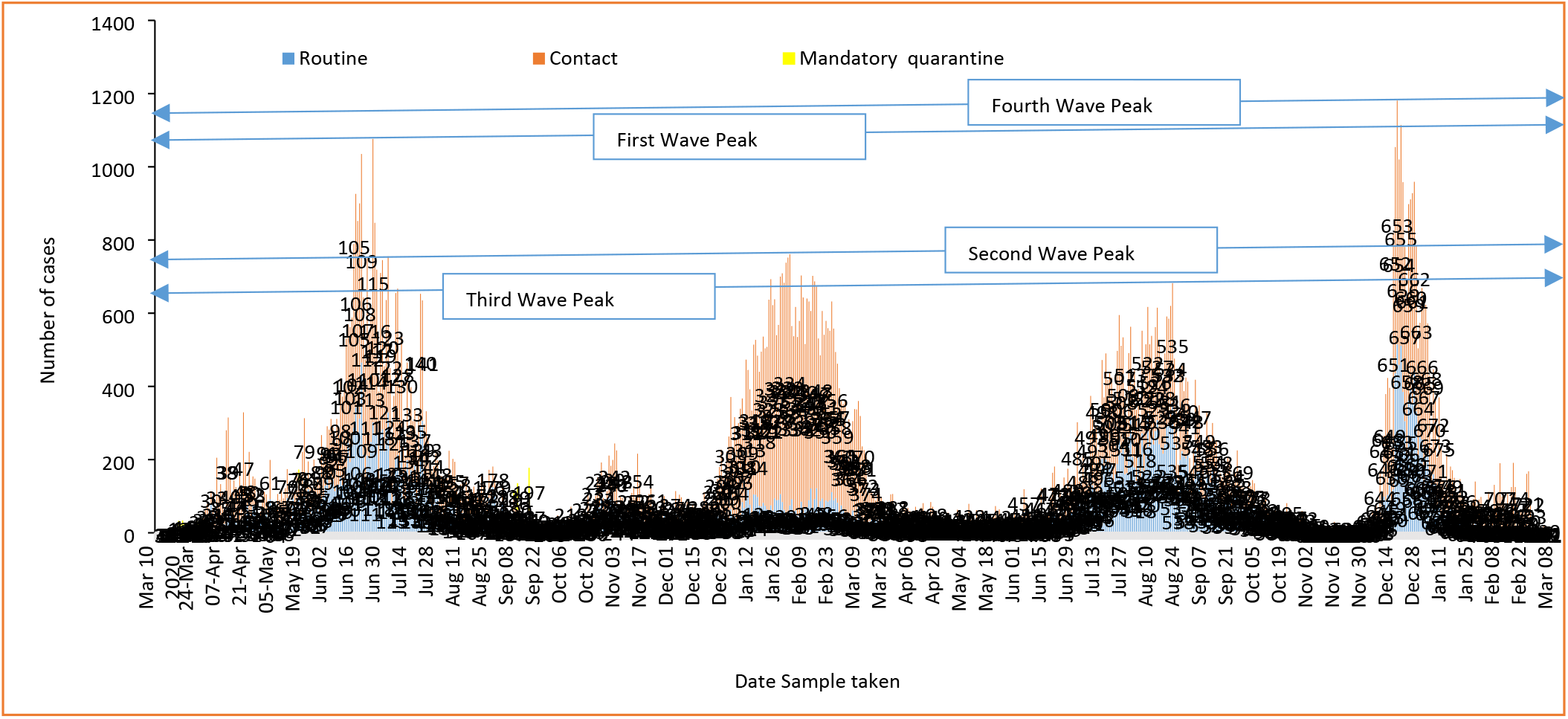
Epidemic waves and distribution of COVID-19 confirmed cases in Ghana, March 2020 – March 2022

**Table 1:** Cumulative COVID-19 confirmed cases, recoveries/discharges, active cases and related mortalities in Ghana, March 2020 - March 2022

| Region | Cases | Proportion (%) | Recovered/Discharged | Proportion Recovered/Discharged | Active Cases | Proportion (%) | Deaths | Case Fatality Rate (%) |
| --- | --- | --- | --- | --- | --- | --- | --- | --- |
| Greater Accra | 90,569 | 56.34 | 90,217 | 99.61 | 35 | 39.33 | 317 | 0.35 |
| Ashanti | 22,298 | 13.87 | 21,903 | 98.23 | 6 | 6.74 | 389 | 1.74 |
| Western | 8,323 | 5.18 | 8,246 | 99.07 | 2 | 2.25 | 75 | 0.90 |
| Eastern | 7,034 | 4.38 | 6,885 | 97.88 | 1 | 1.12 | 148 | 2.10 |
| Central | 5,402 | 3.36 | 5,353 | 99.09 | 0 | 0.00 | 49 | 0.91 |
| Volta | 5,973 | 3.72 | 5,880 | 98.44 | 3 | 3.37 | 90 | 1.51 |
| Northern | 1,863 | 1.16 | 1,831 | 98.28 | 0 | 0.00 | 32 | 1.72 |
| Bono East | 2,966 | 1.84 | 2,892 | 97.51 | 0 | 0.00 | 74 | 2.49 |
| Upper East | 1,734 | 1.08 | 1,672 | 96.42 | 1 | 1.12 | 61 | 3.52 |
| Bono | 2,332 | 1.45 | 2,236 | 95.88 | 0 | 0.00 | 96 | 4.12 |
| Western North | 1,112 | 0.69 | 1,099 | 98.83 | 0 | 0.00 | 13 | 1.17 |
| Ahafo | 1,136 | 0.71 | 1,101 | 96.92 | 1 | 1.12 | 34 | 2.99 |
| Upper West | 895 | 0.56 | 852 | 95.20 | 0 | 0.00 | 43 | 4.80 |
| Oti | 930 | 0.58 | 921 | 99.03 | 0 | 0.00 | 9 | 0.97 |
| North East | 384 | 0.24 | 373 | 97.14 | 0 | 0.00 | 11 | 2.86 |
| Savannah | 291 | 0.18 | 287 | 98.63 | 0 | 0.00 | 4 | 1.37 |
| International travellers (KIA) | 7,519 | 4.68 | 7,479 | 99.47 | 40 | 44.94 | 0 | 0.00 |
| **Total** | **160,761** | **100** | **159,227** | **99.05** | **89** | **100.00** | **1,445** | **0.90** |

As of 12 March 2022, the active COVID-19 cases were 89 with most (40/89) of the cases detected at the Kotoka International Airport (Table 1). Nine administrative regions did not have active cases as of 12 March 2022.

The median daily number of cases in the first minor wave was 85 cases (range: 58 – 353). In the first major wave, the median daily cases were 498 (range: 147 – 1,076). The median confirmed cases in the second minor wave were 115 (range: 41 – 311) per day while the median confirmed cases for the second major wave was 459 (range: 102 – 798) per day. In the third major wave, the median daily new cases were 276 ranging from 35 to 688 cases per day while the fourth wave recorded 166 daily median cases (range: 40 – 1,324). The fourth wave was the shortest in duration but recorded the highest peak of 1,324 cases per day (Figure 1).

A total of 1,445 COVID-19 related deaths were reported as of 12 March 2022. The Ashanti (26.92%), Greater Accra (21.94%) and the Eastern (10.24%) regions accounted for 59.10% (854/1445) of COVID-19 related deaths. The national average Case Fatality Rate (CFR) was 0.90%. The case Fatality ranged from 0.35% in the Greater Accra Region to 4.80% in the Upper West Region (Table 1).

### 4.1 Distribution of Cases

The Greater Accra Region accounted for 56.34% (90569/160761) of all the cases reported as of 12 March 2022 (Table 1). Twelve of the 16 regions had reported more than 1,000 cases of COVID-19 (Figure 2). Similarly, the Greater Accra Region had the highest number of active cases among the regions accounting for 39.33% of the total active cases as of 12 March 2022.

**Figure 2:**
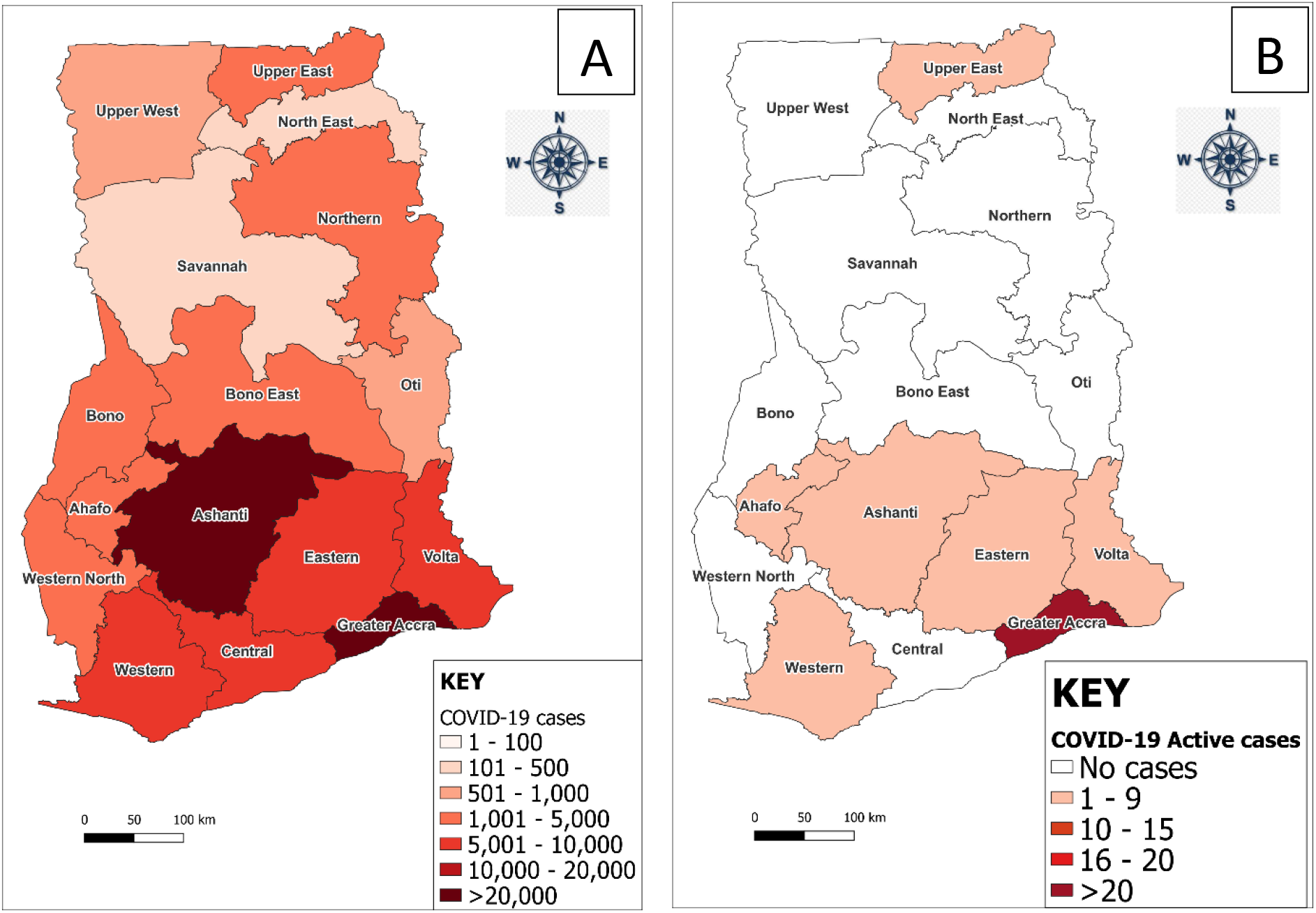
Regional distribution of cumulative confirmed cases (A) and active cases (B) in Ghana, March 2020 - March 2022

### 4.4 Measures taken by government to address COVID-19 pandemic in Ghana

The coordination structures established spanned from the Presidency to the technical operation level. The highest coordination level was the Inter-Ministerial Committee which was chaired by the President of the Republic of Ghana (Figure 3). The technical and operational areas focused on the six identified thematic areas including surveillance, case management, laboratory and testing, Risk Communication and Community Engagement, logistics/procurement and resource mobilization (Figure 4).

**Figure 3:**
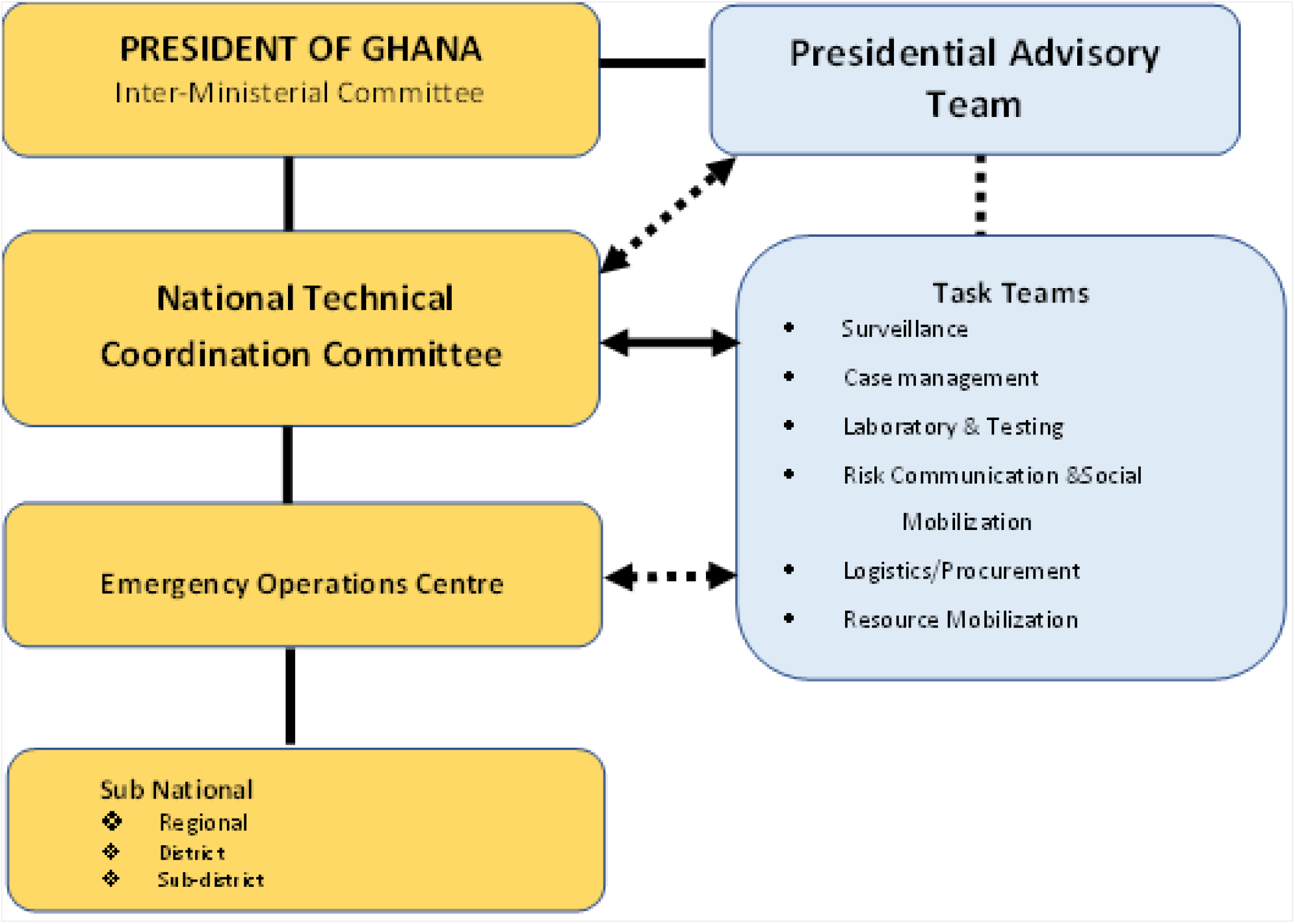
Governance and Coordination Structure for COVID-19 response in Ghana (Source: (5)

**Figure 4:**
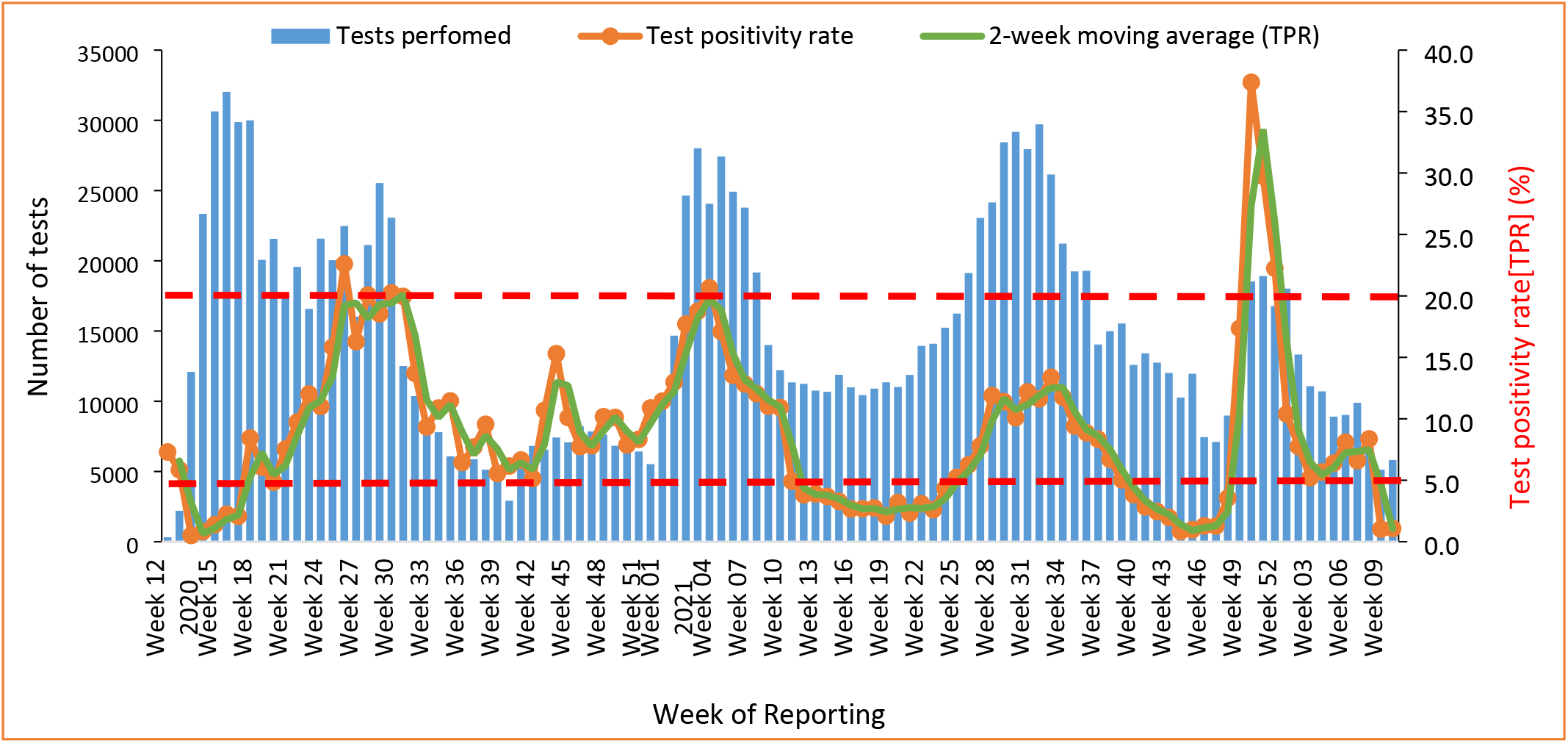
Weekly COVID-19 tests from routine and contact testing in Ghana, March 2020 - March 2022

#### 4.4.1 Declaration of COVID-19 in Ghana

The Government of Ghana, through the Minster of Health, announced the confirmation of two cases of COVID-19 in Ghana on 12 March 2020 (13). This announcement initiated the first contact tracing for the confirmed cases. All contacts underwent mandatory quarantine for at least 14 days following the last date of exposure (2,8). Testing of contacts on mandatory quarantine resulted in the detection of additional cases.

#### 4.4.2 Closure of schools and ban on public gatherings

In line with the Public Health Act (14), the President announced the closure of all schools and a temporary ban on public gatherings effective 16 March 2020 (Figure 6). These measures were to ensure public safety and limit the rate of transmission of the disease.

Large gatherings for funerals were banned. Instead, private burials with a maximum of 25 attendees were allowed. All political rallies, sporting events, restaurants, religious activities including church services and mosques were also banned for initial 4 weeks. The Ministry of Transport engaged transport unions to ensure that enhanced hygienic conditions such as handwashing facilities, hand sanitizers, use of face masks by passengers and drivers and physical spacing in vehicles are observed.

#### 4.4.2 Closure of international borders

All borders (seaports, airports and ground crossings) were closed to human traffic effective 22 March 2020 for a period of 2 weeks subject to reviews. Only conveyances carrying cargo were allowed into the country. Additionally, only returning Ghanaians and foreign nationals with Ghanaian residence permits were allowed into the country during the ban. These returnees were, nonetheless, required to observe a 14-day mandatory quarantine if they showed signs of COVID-19.

#### 4.4.3 Partial Lockdown

On 28 March 2020, the president of the Republic of Ghana, announced a partial lockdown in the Greater Accra Metropolitan Area and Greater Kumasi Metropolitan Area which were observed to be driving community transmission of the disease (15). The lockdown lasted for about 3 weeks from 30 March – 19 April 2020. After lifting the lockdown, the government, in June 2020, further eased the restrictions and allowed the National Identification Authority (NIA) and the Electoral Commission (EC) to conduct registration exercises which were earlier planned but suspended due to COVID-19 (16).

#### 4.4.4 Reopening of Kotoka International Airport

The Kotoka International Airport was officially reopened to passengers for international travel on 01 September 2020 (17) after about 6 months of closure. All international travellers (arriving and departing) were required to strictly adhere to the COVID-19 protocols during the flights and at the airport. Arriving passengers were required to possess a negative PCR test done not more than 72 hours prior to departure as well as a mandatory COVID-19 test (rapid antigen-based test) on arrival at the airport (18). All passengers who tested positive on arrival were sent to the designated hospital for further assessment and management (including mandatory isolation).

#### 4.4.5 Resumption of Contact Sports

In his 17^th^ Nation’s address on measures taken to address the COVID-19 pandemic in Ghana, the President of Ghana lifted the ban on contact sports. Spectators were not allowed at the training centres and only 25% seating capacity at the stadia was allowed to ensure physical distancing. All sports men and women in camp were to be tested regularly.

#### 4.4.6 Reopening of basic schools

The President announced reopening of pre and basic schools The reopening took effect from 18 January 2021. Prior to the reopening, schools were mapped to health facilities for regular monitoring and supervision of adherence to COVID-19 protocols and general health status of school children and staff. The school mapping was also to ensure prompt isolation and treatment of cases detected among school children. Schools were fumigated before reopening. School children and staff were required to strictly adhere to the COVID-19 protocols (19).

#### 4.4.7 Re-imposition of restrictions

Following a resurgence of COVID-19 cases in late December 2020 and in January 2021, the President reimposed some restrictions. Public gatherings such as funerals, weddings, concerts, theatrical performances, and parties were again banned effective 31 January 2021. This event was preceded by two major events in the country. These were National Elections (early December 2020) and Christmas and New Year festivities.

#### 4.4.8 COVID-19 vaccine rollout

Ghana was the first country to receive COVID-19 vaccines through the COVAX facility – a global mechanism to ensure the equitable distribution of COVID-19 vaccines. The vaccination rollout was launched on 01^t^ March 2021 followed by a phased sub-national vaccination campaign. The first phase covered 43 high-risk districts in three regions (Ashanti, Greater Accra and Central). The initial phase of the vaccine rollout targeted a category of persons designated as most at risk for COVID-19 infection and severity of the disease. These included healthcare workers, frontline security personnel, and persons with known underlying medical conditions, persons 60 years and above, frontline members of the Executive, Legislature and Judiciary, and teachers over 50 years of age.

As of 13 March 2022, Ghana had received 27,098,550 doses of COVID-19 vaccinations of which 12,637,214 doses (46.6%) have been administered (20). Ghana targeted to vaccinate at least 70% of the population (approximately 22,859,226). Approximately, 38.5% (8,793,591) persons have received at least one dose while 21.3% (4,886,214) persons had completed the primary vaccination series (fully vaccinated) by 13 March 2023 (20).

### 4.2 Laboratory Testing and levels of community transmission

As of 12 March 2022, a total of 2,376,530 (76,774 tests per million population) tests were conducted with 160,761 testing positive (test positivity rate=6.8%). Of the cumulative tests, 20.21% (480,285) were from contact tracing, 44.17% (1,049,612) from routine surveillance testing and 35.62% (846,633) through testing of international arriving passengers at the Kotoka International Airport (KIA).

The median daily test during the first major wave was 3,091 (range: 809 – 4,641) and a median test positivity of 18.24 % (range: 3.70% - 76.52). During the second minor wave, the median daily test was 990 (range: 567 – 2,477) and the median test positivity rate was 9.93% (range: 2.64 – 26.32). In the second major wave which lasted from first week of January 2021 to the second week of March 2021, the median daily test was 3,046 (range: 1,132 – 4,653) with a median test positivity rate of 16.30% (range: 9.60 – 24.06). The test positivity rate ranged from 0.5% in Week 14 of 2020 to 37.4% in Week 51 of 2021. The highest positivity rate was reported in the fourth major wave (Figure 4). The level of community transmission was high for most parts of the major transmission waves and very high for most weeks of the first transmission week (Figure 4). The fourth major transmission wave recorded the shortest period for which community transmission was very high.

### 4.3 Effective (R_1_) Reproduction number

The highest R_t_ recorded was 8.77 (95% CI: 7.29 – 10.44) on 26 March 2020 (Figure 5). The reproduction rate declined to <1 (0.99, 95% CI: 0.81 – 1.19) for the first time on 31 March 2021. It subsequently rose >1 and spun for 5 weeks (07 April – 15 May 2020) and declined thereafter for another one week.

**Figure 5:**
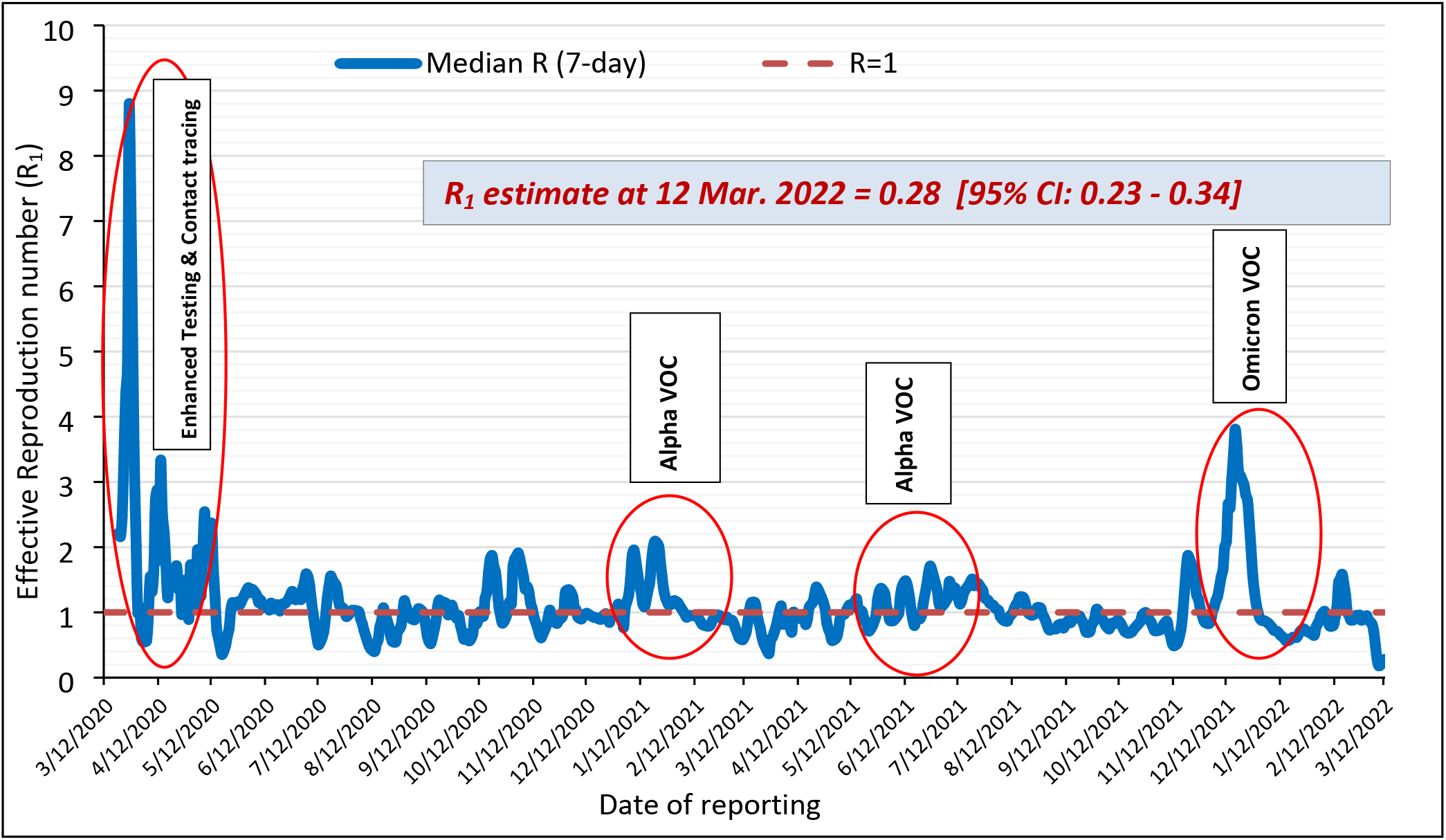
Effective Reproduction rate estimates of COVID-19 in Ghana, March 2020 - March 2022

**Figure 6:**
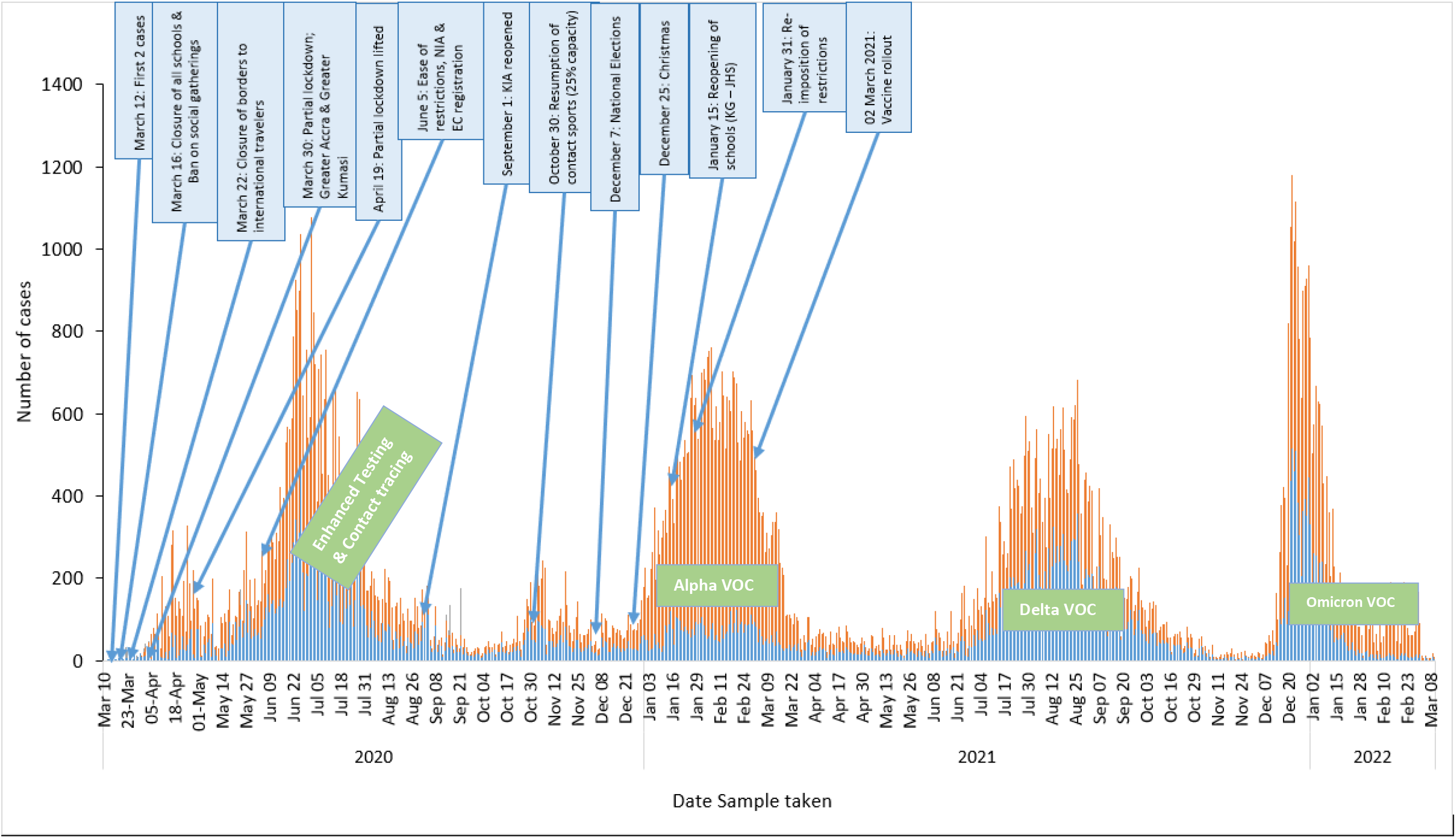
Timelines of measures taken by government in response to the COVID-19 pandemic in Ghana, March 2020 - March 2022

The longest sustained effective reproduction number above 1 spanned approximately 7 weeks (23 May 2020 – 08 July 2020). The highest R_t_ after the Christmas festivities was 2.11 (95% CI: 2.04 – 2.18) on 21 January 2021. Also, the longest period for which R_t_>1 was recorded was for 47 days from 23 May 2020 to 08 July 2020 (Figure 5). The R_t_ on 12 March 2022 was 0.28 (95% CI: 0.23 – 0.34). The highest R_t_ in the fourth wave (R_t_= 3.81 [95% CI: 3.64 – 3.98]) was recorded on 17 December 2021 (Figure 5).

## Discussion

This study examined the trend of COVID-19 pandemic in Ghana in the first two years, described some measures undertaken in the response and examined the level of community transmission in the wake of control measures adopted and other activities which might have fuelled the transmission.

The Greater Accra Region has been the epicentre of COVID-19 in Ghana. Aside the dense population, the region is home to a lot of industries along the coastal enclave and big markets in the metropolis which may be a conduit for COVID-19 infection amidst poor adherence to preventive protocols (21). Despite increased awareness of people on COVID-19, knowledge alone did not translate into adherence to preventive measures (22). This therefore calls for sustained public engagements for attitudinal change.

Ghana recorded one of the lowest Case Fatality Rate CFR) for COVID-19 compared with many countries in the Sub-Saharan Africa (23). The CFR at the end of the fourth wave was similar to what was reported in the Democratic Republic of Congo (24). Ghana has a relatively young population (25) and less than 10% of COVID-19 infections were >60 years (26) which might explain the low case fatality rate. At the sub-national level, there was, however, an observed higher case fatality rate in regions in the northern part of the country than in the middle and southern belt. This could be due to the disproportionate distribution of both human resources and well-equipped health facilities (27) to handle severe and critical COVID-19 cases in the country. The major state-of-the-art Infectious Disease Centre is in the Greater Accra region which may explain the lowest case fatality rate recorded in the region.

The institution of strict restrictions in the early stages of pandemic in Ghana slowed transmission of the disease for the first one month. The initial spike observed in April 2020 could be attributed to the enhanced contact listing and testing (8) accounting for most asymptomatic cases (26). The first major wave recorded in Ghana was preceded by the ease of some restrictions including the lifting of the partial lockdown and the allowance of mass gatherings for the National Identification Authority (NIA) and the Electoral Commission (EC) to register citizens for various exercises. The significant nonadherence to the use of face mask in some major cities including in public transport (28) could fuel transmission. In a study assessing the effects of outbreak response and interventions on COVID-19 waves in Africa, Ilesanmi and colleagues observed that the stringent implementation of public health and social measures including border closures and screening of international travellers effectively decreased the number of COVID-19 cases recorded (29). This finding reinforces the importance of Public Health and Social Measures as an effective means in preventing the spread of COVID-19 (30,31).

Enhanced contact tracing coupled with an innovative approach of sample pooling made Ghana the country with the highest testing rate per population (approximately 173 tests per million population in Africa as of April 2020 (32). The sample pooling strategy reduced the turn-out time for laboratories, cleared huge sample backlogs and rapidly linked cases to care (Asante et al., 2021). Improving turn-around time essentially could contribute to slowing transmission as confirmed cases are more willing to isolate when they know their test results (33).

Four distinct major waves of COVID-19 were observed in Ghana. These waves were consistent with the periods observed in many other countries worldwide (24,34). Interestingly, the major waves were characterized by mutant variants of concern which had different impact on disease transmissibility and degree of severity on the infected persons (35). The Alpha variant was initially detected in the United Kingdom in September 2020 (35) and from January - March 2021 (second wave), it became the predominant variant of concern in Ghana (36). This might have been exacerbated by the harsh harmattan winds (37) which characterize this period.

The third major wave in Ghana was predominantly characterized by the Delta VOC (36). The Delta variant which was first detected in India rapidly spread across many continents within a short period (38). The VOC was first detected in Ghana in May 2021 (36) and by mid-July, the variant was detected among community members without travel history (39). The variant was known for its higher transmissibility, increased hospitalization of infected persons, higher viral loads, and reduced response to existing COVID-19 vaccines (38). Though the number of cases reported in the third wave were less than those of the second wave, hospitalization increased putting a strain on the health infrastructure especially the treatment facilities.

The fourth wave was primarily characterized by the Omicron Variant (40,41). The omicron variant was noted for its increased transmissibility, and ability to escape neutralizing antibodies of already vaccinated persons largely due to the several (32) spike proteins it possessed (40). Reinfection of previously infected individuals was highly possible since previous vaccination did not offer adequate protection against the variant (36,38,41).

One of the key interventions by many governments in response to the COVID-19 pandemic in the early stages was the imposition of lockdowns. This intervention was not only to contain the disease and break the transmission chain but to also buy time and to understand the disease dynamics (1). In Ghana’s context, the lifting of lockdown was guided more by economic rather than scientific evidence (42). The lockdown was hastily lifted, without adequate communication and transparency (1) and with little recourse to established guidelines (3,43).

## Study limitations

This study used secondary data published on the Ghana Health Service COVID-19 website. The data had no sex and age disaggregation. Therefore, analysis on the impact of the disease on children and gender dynamics could not be done in this study. In a previous study during the early stages of the pandemic in the country (26), evidence suggested that more males than females were infected and died. As safe vaccines for children <15 years are yet to be made available in Ghana, there is the need to understand the impact of the disease in such age groups.

The cumulative number of cases at a given time may also include individuals who might have been re-infected since the data did not disaggregate reinfections. Some individuals might have been re-infected several times but still reported as individual case counts. This might overestimate the number of infected cases at a given time. However, the impact of cases on the health system does not differ significantly irrespective of whether they are first time cases or reinfections.

## Conclusions

The pattern of COVID-19 waves in Ghana was similar to the global trend. The waves were characteristically driven by variants of concerns. Vaccination might not have contributed to a reduction in the number of new infections but seems to have positive impact on disease severity and mortality.

The whole-of-government and whole-of-society approach in the COVID-19 response provided direction for the response. The measures undertaken by government in COVID-19 showed mixed impact on COVID-19 transmission. High community transmission was heralded by the detection of a new variant of concern. The level of community transmission was highest in the fourth wave. COVID-19 might stay with humanity longer than initially anticipated. Even though the control measures largely remain unchanged, a review of these measures as new evidence becomes available is essential to ensure compliance. There is the need to enhance vaccination drive especially for populations with increased risk for disease severity, increase laboratory capacity for genomic surveillance and monitor resurgence. These measures may reduce the risk of the emergence of new variants as well as minimize the risk of severity associated with new variants of concern.

## Data Availability

Dataset used for this manuscript is public available. All extracted data and analysis can be made available on request

https://ghs.gov.gh/covid19/archive.php#

